# Sensing and decoding the neural drive to paralyzed muscles during attempted movements of a person with tetraplegia using a sleeve array

**DOI:** 10.1101/2021.02.24.21250962

**Authors:** Jordyn E. Ting, Alessandro Del Vecchio, Devapratim Sarma, Samuel C. Colachis, Nicholas V. Annetta, Jennifer L. Collinger, Dario Farina, Douglas J. Weber

**Affiliations:** Rehab Neural Engineering Labs, University of Pittsburgh, Pittsburgh, PA, 15213, USA; Department of Bioengineering, University of Pittsburgh, Pittsburgh, PA, 15213, USA; Center for the Neural Basis of Cognition, Pittsburgh, PA, 15213, USA; Department of Artificial Intelligence in Biomedical Engineering, Friedrich-Alexander University, Erlangen-Nürnberg, Erlangen, 91052, Germany; Department of Mechanical Engineering, Carnegie Mellon University, Pittsburgh, PA 15213; Neuroscience Institute, Carnegie Mellon University, Pittsburgh, PA 15213; Medical Devices and Neuromodulation Group, Battelle Memorial Institute, Columbus, OH, 43201, USA; Department of Physical Medicine and Rehabilitation, University of Pittsburgh, Pittsburgh, PA, 15213, USA; Human Engineering Research Laboratories, VA Center of Excellence, Department of Veterans Affairs, Pittsburgh, PA, 15206, USA; Department of Biomedical Engineering, Carnegie Mellon University, Pittsburgh, PA 15213; Department of Bioengineering, Imperial College London, London, UK

## Abstract

Motor neurons in the brain and spinal cord convey information about motor intent that can be extracted and interpreted to control assistive devices, such as computers, wheelchairs, and robotic manipulators. However, most methods for measuring the firing activity of single neurons rely on implanted microelectrodes. Although intracortical brain-computer interfaces (BCIs) have been shown to be safe and effective, the requirement for surgery poses a barrier to widespread use. Here, we demonstrate that a wearable sensor array can detect residual motor unit activity in paralyzed muscles after severe cervical spinal cord injury (SCI). Despite generating no observable hand movement, volitional recruitment of motor neurons below the level of injury was observed across attempted movements of individual fingers and overt wrist and elbow movements. Subgroups of motor units were coactive during flexion or extension phases of the task. Single digit movement intentions were classified offline from the EMG power (RMS) or motor unit firing rates with median classification accuracies >75% in both cases. Simulated online control of a virtual hand was performed with a binary classifier to test feasibility of real time extraction and decoding of motor units. The online decomposition algorithm extracted motor units in 1.2 ms, and the firing rates predicted the correct digit motion 88 ± 24% of the time. This study provides the first demonstration of a wearable interface for recording and decoding firing rates of motor neurons below the level of injury in a person with tetraplegia after motor complete SCI.

**Significance Statement:** A wearable electrode array and machine learning methods were used to record and decode myoelectric signals and motor unit firing in paralyzed muscles of a person with motor complete tetraplegia. Motor unit action potentials were extracted from myoelectric signals during attempted movements of the fingers and voluntary movements of the wrist and elbow. The patterns of EMG and motor unit firing rates were highly task-specific, even in the absence of visible motion in the limb, enabling accurate classification of attempted movements of single digits. These results demonstrate the potential to create a wearable sensor for determining movement intentions from spared motor neurons, which may enable people with severe tetraplegia to control assistive devices such as computers, wheelchairs, and robotic manipulators.

## Introduction

The hand is arguably the most complex apparatus of the motor system, possessing up to 24 kinematic degrees of freedom making it capable of a wide range of tasks essential for accomplishing activities of daily living (ADLs) (1). Thus, the loss of hand function, especially when bilateral, has a profound impact on a person’s ability to live and thrive independently (2), making recovery of hand function a top priority for people with tetraplegia after spinal cord injury (SCI) (3, 4). Functional electrical stimulation (FES) and powered exoskeletons may be used to reanimate the paralyzed hand, but these and other assistive devices require a way for users to express movement intentions. To date, intracortical brain-computer interfaces (BCIs) have demonstrated the greatest potential for enabling people with tetraplegia to control complex, multifunctional movements of the hand and arm (5–8). By comparison, few groups have explored the potential of myoelectric control in people with tetraplegia, although recent reports provide promising evidence demonstrating the feasibility of obtaining myoelectric control signals from muscles weakened or paralyzed by SCI (9–12). Here, we show that task-specific patterns of EMG and motor unit firing can be detected in muscles below the level of SCI and decoded to predict attempted movements of individual digits in the paralyzed hand. These findings demonstrate the potential of using a wearable sensor array to control assistive devices for restoring voluntary, multi-degree of freedom functions in the hand.

The presence of volitional myoelectric activity in muscles that are diagnosed clinically as completely paralyzed indicates that at least part of the corticospinal tract remains intact. Such SCIs have been termed *discomplete*, a term encompassing spinal cord damage that presents clinically as motor complete (no voluntary motor function) but retaining some detectable muscle activity in subclinical testing (13). While the remaining muscle activity is not sufficient to generate overt movements, even weak myoelectric signals may be used as a source of voluntary control signals for operating assistive devices, even after complete SCI (9). Surface EMG signals recorded from the hands and forearms of individuals with both motor incomplete and motor complete SCIs have been used to decode hand grasp intentions (9–11, 14). Lu et al. demonstrated real-time control of an exoskeleton, using both bipolar EMG and force measurements, to assist 3 full-range hand movements in participants with tetraplegia (9). However, bipolar EMG in these muscles are inadequate for detecting sufficient information to decode user intent, resulting in imprecise and inaccurate control signals, especially for subjects with more severe levels of impairment and for the recovery of complex dexterous movements of the hand. Motor unit activity, however, can provide a more direct representation of motor commands, since the motor unit action potentials are directly coupled to the output of spinal motor neurons.

Myoelectric signals can be detected easily with epidermal sensors that can be integrated into the fabric of form-fitting garments to create a wearable sensing interface. For example, myoelectric sensors are used routinely for controlling state-of-the-art powered prosthetic hands with arrays of electrodes mounted in the liner of the prosthesis (15). However, myoelectric signals, if present, will be weak and sparse in muscles that are severely paralyzed (12). In this case, to achieve accurate and precise detection and classification of myoelectric activity requires a sensing array that provides broad coverage of the paralyzed limb and a method for detecting features of the EMG activity that are linked robustly to motor intent.

In the present study, we addressed these challenges in two ways. First, we developed a novel myoelectric sensor array containing 150 electrodes embedded in a stretchable fabric to measure EMG from the entire surface of the forearm (Fig. 1A-B) (14). This sleeve array can be positioned quickly on the arm without any specialized technical expertise. The sleeve measured EMG in the forearm of a participant with tetraplegia, revealing weak but task-specific patterns of myoelectric activity. Motor unit decomposition was performed on the EMG to extract discharge timing information for small groups of isolated motor units. Signal quality metrics, including signal to noise ratio for the sleeve EMG were similar to that of a commercial high-density EMG (HDEMG) system tested on the same participant. Since the motor unit recordings are a direct measure of the output from spinal motor neurons, their discharge timings provide the highest possible spatial and temporal resolution for measuring motor activity. This information was used for decoding the motor intent during attempted movements of individual digits.

**Figure 1.**
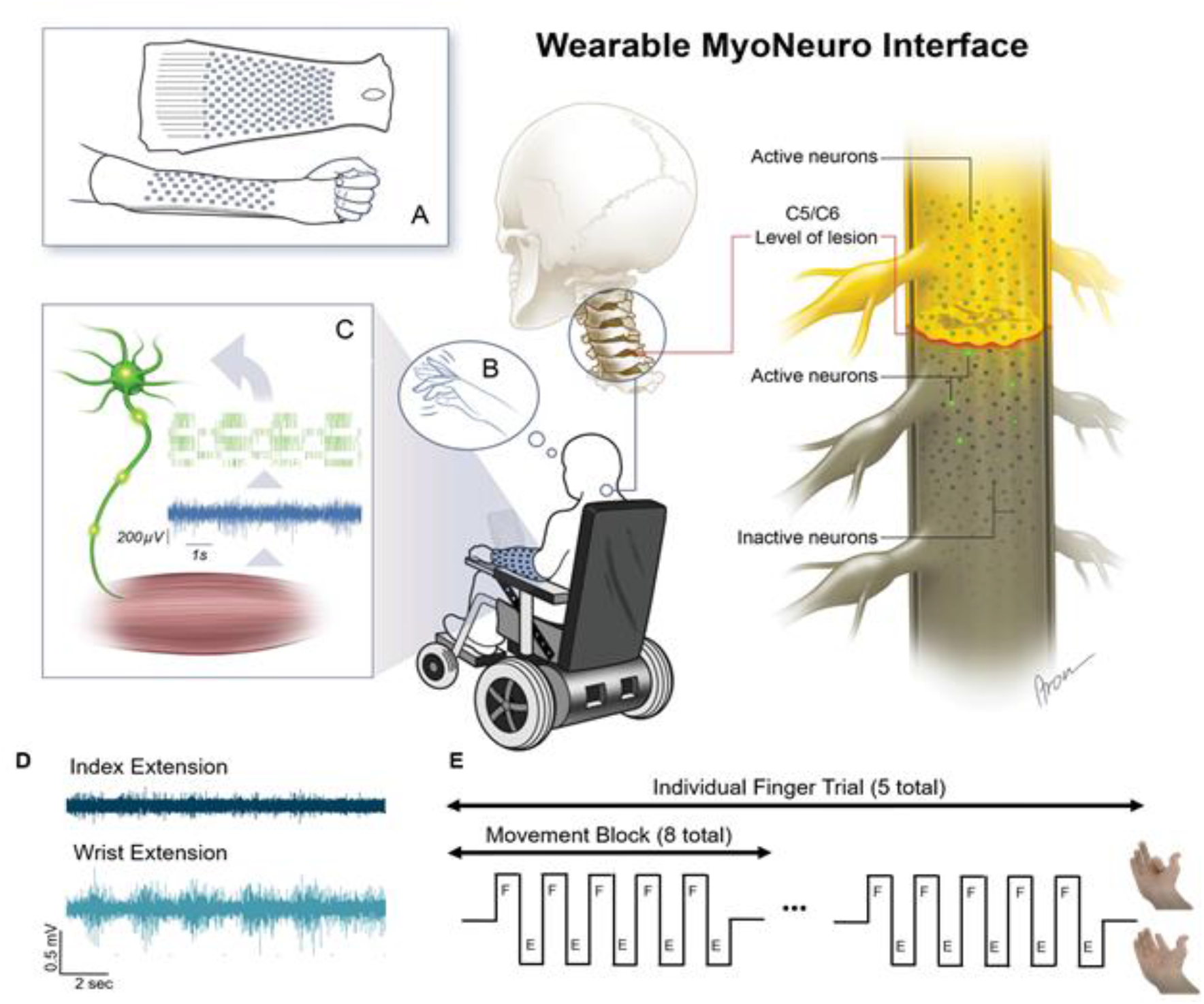
Experimental setup. **A** Custom sleeve array developed to record myoelectric signals from the forearm muscles. **B** The participant was instructed to attempt to flex and extend his fingers but was unable to produce any visible movement. **C** Myoelectric signals were detected in the forearm muscles and were decomposed into the activity of individual, active motor units. **D** EMG signals were weaker during attempted index extensions than during voluntary wrist extensions. **E** The experiment consisted of eight “movement blocks” comprising five alternating flexion and extension movements of individual fingers, wrist, and elbow. An example of the visual cues for index flexion and extension are shown.

This study provides the first demonstration of a direct interface with central nervous system (CNS) neurons in a person with a motor complete SCI using a wearable sensor array. Motor unit action potentials from 20 motor units were isolated during attempted movements of the fingers, which are completely paralyzed, and movements of the wrist and elbow that the participant could perform overtly. Despite a complete absence of visible motion in the digits, the motor units exhibited clear and task-specific increases in discharge rate during attempted movements of the fingers. The patterns of motor unit firing generated during attempted movements were highly discriminable across tasks, highlighting the potential to create a neural interface for controlling devices that assist recovery of motor function in the paralyzed hand. Moreover, the algorithms for detecting and decoding motor unit firing rates are computationally efficient and can be implemented in real-time for online control of assistive devices.

## Results

### Myoelectric Activity in Intact and Paralyzed Muscles

A 150-channel sleeve electrode array was used to measure myoelectric activity across the forearm of an individual with tetraplegia during attempted finger movements and overt wrist and arm movements (Fig. 1). The participant was able to flex and extend his wrist and elbow but unable to produce detectable movement in any digit as confirmed by videography. A Manual Muscle Test (MMT) was performed to evaluate the strength and function of the participant’s wrist flexors and extensors and finger flexors and extensors (16). The wrist extensors scored 4-, indicating that the participant could move through extension when slight to moderate pressure was applied. The wrist flexors, finger flexors, and finger extensors scored 0, meaning that no contractions were visible or palpable in the muscles. Two examples of bandpass-filtered surface EMG are shown to illustrate the myoelectric signals recorded during attempted movements of the index finger and overt movements of the wrist (Fig. 1D). While the EMG amplitude was lower and sparser during attempted movements of the index finger, the myoelectric signals were clearly detectable above the baseline in both task conditions.

The heatmaps in Fig. 2 illustrate the EMG power root-mean-square (RMS) across the 10 x 15 grid of electrodes (Fig. 1A) during the movement block when the subject was instructed to attempt alternating flexion and extension movements (Fig. 1E). Distinct, task-specific zones of EMG were detected on both sides of the forearm, but the myoelectric signals were stronger and more widespread on the extensor side. The largest EMG was measured during overt movements of the wrist; note the difference in scale between the wrist and digits. Although no motion was produced in any of the digits, attempted movement of the thumb, index, and middle fingers produced clear patches of EMG on the extensor side and weaker activation on the flexor side. Attempted movement of the ring and pinky fingers produced weak activity on the extensor side, while myoelectric signals on the flexor side were barely visible.

**Figure 2.**
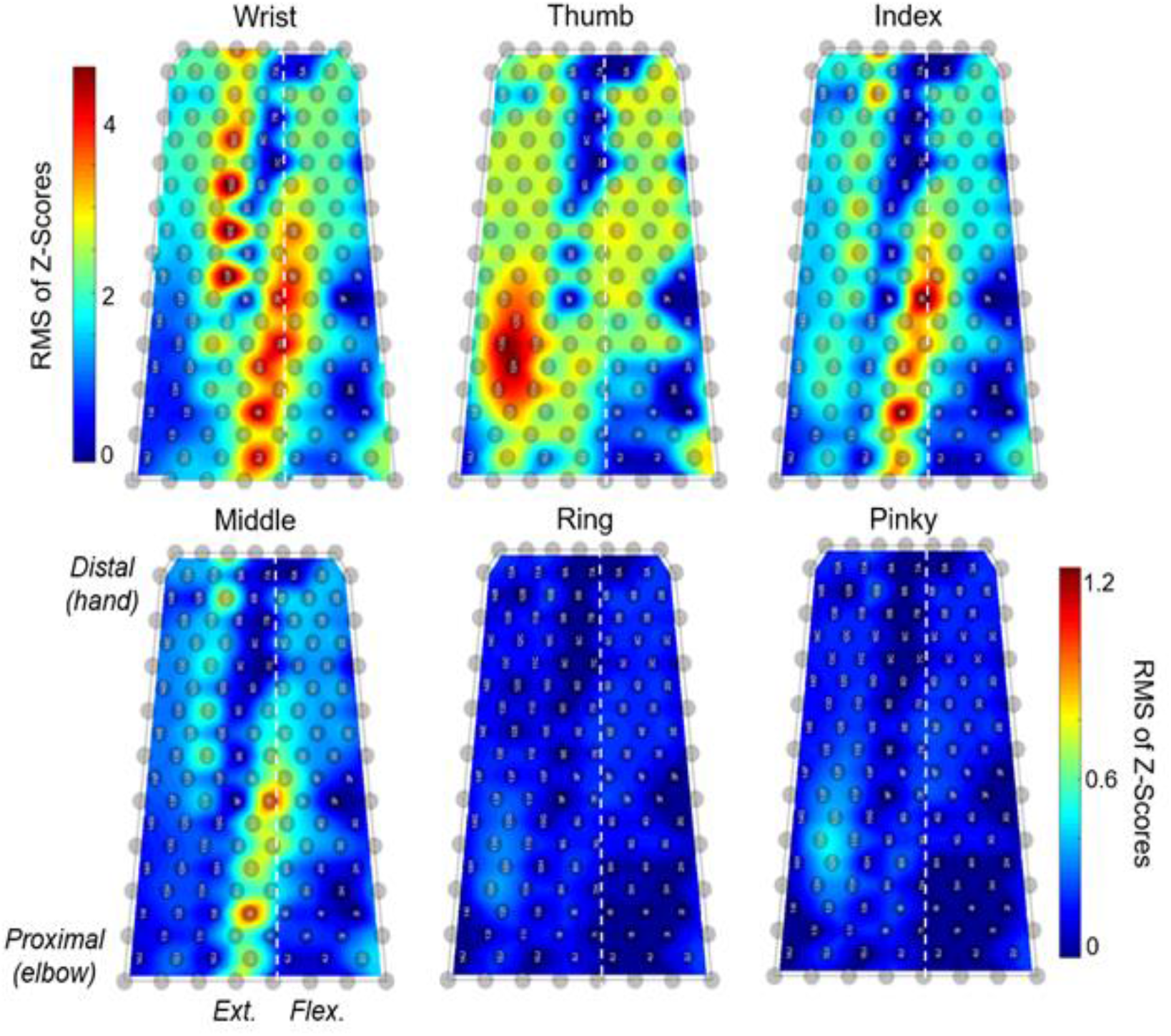
Heatmaps of monopolar EMG power during overt and attempted flexion-extension movements. EMG power during the movement block was measured as the root-mean-square (RMS) of the bandpass-filtered and z-score transformed EMG for each electrode (gray circles) in the sleeve array. The color bar in the top row corresponds to the heatmap for the wrist, and the color bar in the bottom row represents the scale for the single digit heatmaps. The left and right edges of the heatmap correspond to the border between the dorsal and ventral sides of the forearm on the ulnar side. The vertical dashed line in each plot represents the approximate location of the border between the dorsal and ventral sides on the radial side of the forearm. The extensor and flexor muscle groups are located to the left and right of the dashed line, respectively.

### Detection of Motor Unit Action Potentials

Motor unit decomposition (17) was performed to identify motor unit action potentials (MUAPs) from surface EMG recorded with the sleeve array. The validity of motor unit decomposition was assessed via two-dimensional cross-correlation of the MUAP waveforms, taken between random firings of the motor unit. This analysis yielded high correlation values >0.7, compared to chance level correlations, which ranged between 0.1 and 0.2.

Table 1 shows the number of identified motor neurons across all attempted arm and digit movements. A total of 20 unique motor units were identified. Of these, 14 were active during a single task and 6 were active during two or more tasks. The average number of identified motor neurons across tasks was 3.9 (s.d. = 1.8 units). The wrist extension/flexion task yielded the highest number of detectable motor units (n=7), while only one motor neuron was identified during attempted thumb movements. For all tasks, except the thumb, multiple motor units were detected and all exhibited increased firing rates during movement blocks.

**Table 1.**
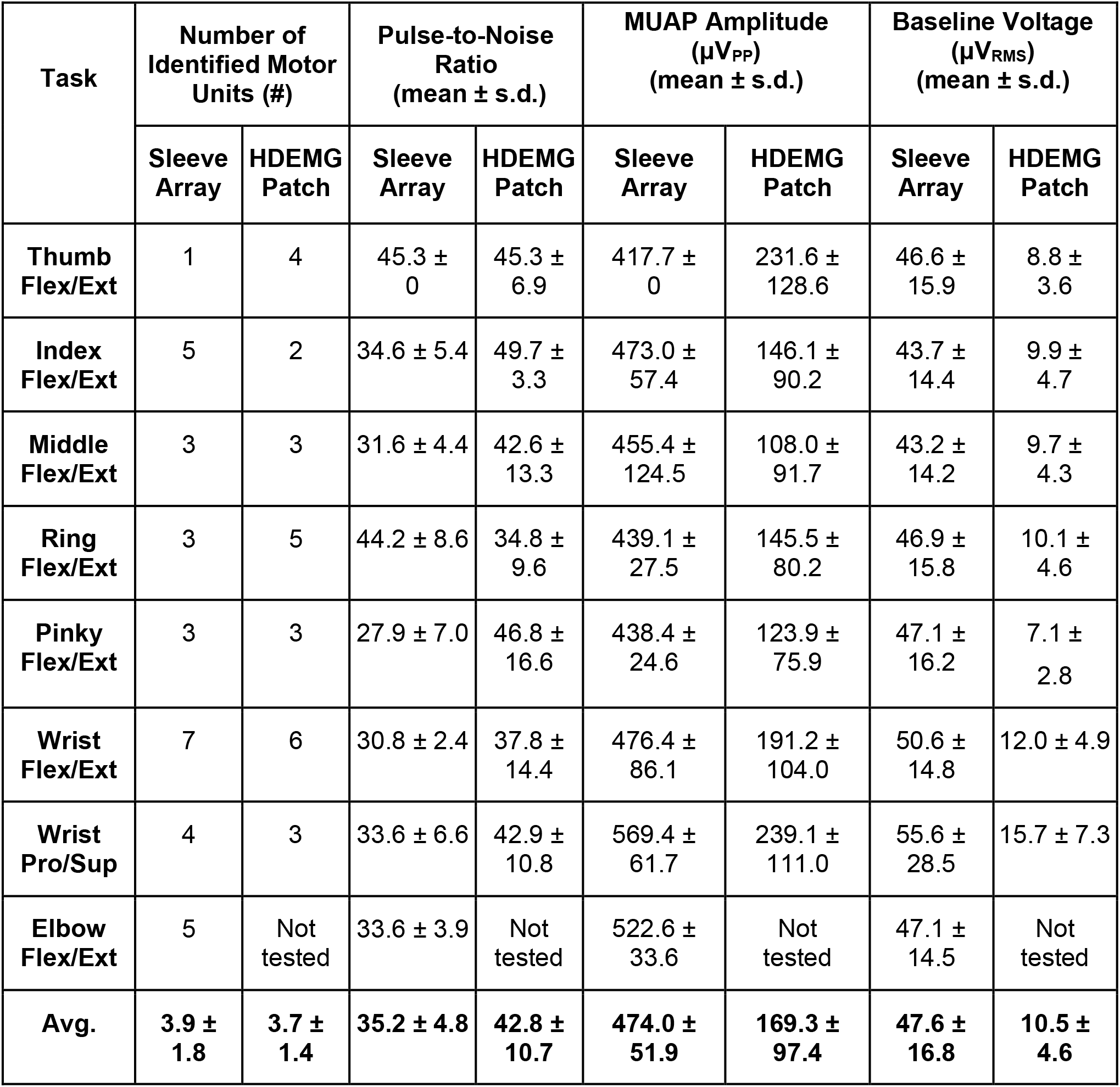
Motor unit, EMG signal, and noise properties for the sleeve and HDEMG patch arrays. The total number of motor units identified during each task are reported when the sleeve array (150 electrodes) and the HDEMG patch (one 64-channel array on dorsal aspect) were used to record EMG. Only data from the HDEMG patch on the extensor side of the forearm is reported here. The pulse-to-noise ratio (PNR) is an estimate of the signal-to-noise ratio of the extracted motor units. The motor unit action potential (MUAP) amplitude for each unit is taken from the average of all waveforms associated with that unit detected during a particular task. The PNRs and MUAP amplitudes are reported as the mean and standard deviation across the motor units identified during each task. The baseline voltage was sampled during ‘rest’ periods where the participant was instructed to relax his arm. The baseline voltage is reported as the mean and standard deviation of the RMS voltages taken across all electrodes in the sleeve array or patch.

| Task | Number of Identified Motor Units (#) | | Pulse-to-Noise Ratio (mean ± s.d.) | | MUAP Amplitude (μV<sub>PP</sub>) (mean ± s.d.) | | Baseline Voltage (μV<sub>RMS</sub>) (mean ± s.d.) | |
| --- | --- | --- | --- | --- | --- | --- | --- | --- |
|  | Sleeve Array | HDEMG Patch | Sleeve Array | HDEMG Patch | Sleeve Array | HDEMG Patch | Sleeve Array | HDEMG Patch |
| **Thumb Flex/Ext** | 1 | 4 | 45.3 ± 0 | 45.3 ± 6.9 | 417.7 ± 0 | 231.6 ± 128.6 | 46.6 ± 15.9 | 8.8 ± 3.6 |
| **Index Flex/Ext** | 5 | 2 | 34.6 ± 5.4 | 49.7 ± 3.3 | 473.0 ± 57.4 | 146.1 ± 90.2 | 43.7 ± 14.4 | 9.9 ± 4.7 |
| **Middle Flex/Ext** | 3 | 3 | 31.6 ± 4.4 | 42.6 ± 13.3 | 455.4 ± 124.5 | 108.0 ± 91.7 | 43.2 ± 14.2 | 9.7 ± 4.3 |
| **Ring Flex/Ext** | 3 | 5 | 44.2 ± 8.6 | 34.8 ± 9.6 | 439.1 ± 27.5 | 145.5 ± 80.2 | 46.9 ± 15.8 | 10.1 ± 4.6 |
| **Pinky Flex/Ext** | 3 | 3 | 27.9 ± 7.0 | 46.8 ± 16.6 | 438.4 ± 24.6 | 123.9 ± 75.9 | 47.1 ± 16.2 | 7.1 ± 2.8 |
| **Wrist Flex/Ext** | 7 | 6 | 30.8 ± 2.4 | 37.8 ± 14.4 | 476.4 ± 86.1 | 191.2 ± 104.0 | 50.6 ± 14.8 | 12.0 ± 4.9 |
| **Wrist Pro/Sup** | 4 | 3 | 33.6 ± 6.6 | 42.9 ± 10.8 | 569.4 ± 61.7 | 239.1 ± 111.0 | 55.6 ± 28.5 | 15.7 ± 7.3 |
| **Elbow Flex/Ext** | 5 | Not tested | 33.6 ± 3.9 | Not tested | 522.6 ± 33.6 | Not tested | 47.1 ± 14.5 | Not tested |
| **Avg.** | **3.9 ± 1.8** | **3.7 ± 1.4** | **35.2 ± 4.8** | **42.8 ± 10.7** | **474.0 ± 51.9** | **169.3 ± 97.4** | **47.6 ± 16.8** | **10.5 ± 4.6** |

The average peak-to-peak amplitude of the MUAPs detected by the sleeve array was 474.0 μV (s.d. = 51.9 μV). The MUAP amplitudes were similar across tasks, although the MUAPs detected during overt movements of the wrist and elbow were slightly larger than the MUAPs detected during attempted movements of the digits (p<0.05). The average pulse-to-noise ratio (PNR), which is a measure of the accuracy of the EMG decomposition (18), was 35.2 ± 4.8 and was invariant across tasks.

The motor unit decomposition procedure isolates MUAP waveforms using a combination of signals recorded on multiple electrodes via blind source separation procedures (17). To visualize the MUAP waveforms as they appear on a single electrode, we used the discharge times for each motor unit to generate a spike-triggered average of the EMG on each electrode in the sleeve array. Figure 3 (A-C) shows three representative MUAP waveforms isolated by spike-triggered averaging.

**Figure 3.**
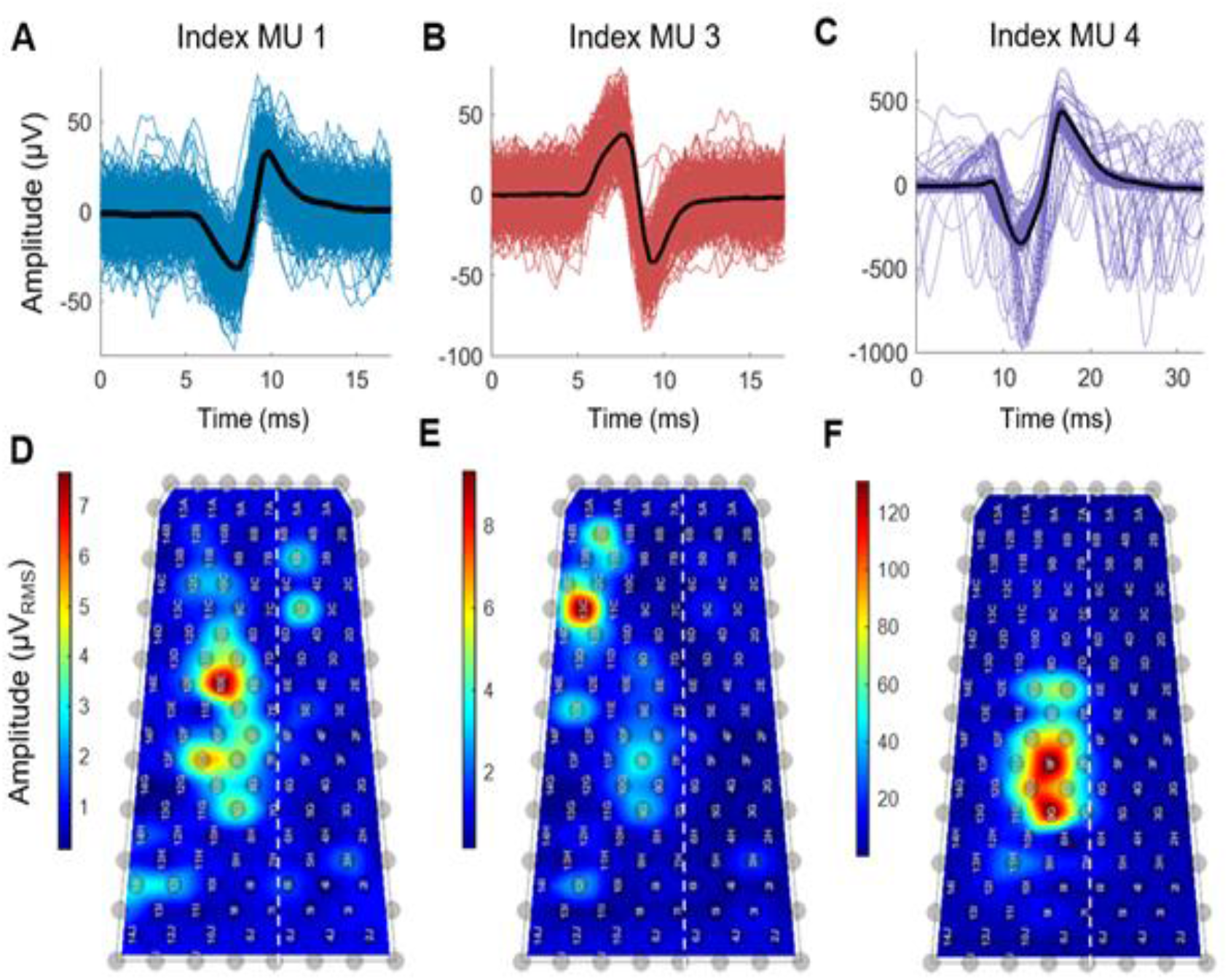
Motor unit action potentials identified after decomposition. **A-C** MUAP waveforms were identified by decomposition during attempted movements of the index finger. Three of five motor units detected during the index finger trial are shown. MUAP waveforms were extracted by spike-triggered averaging the firing instances of each motor neuron. The MUAP waveforms of three representative units are shown with the average waveform of each unit overlaid (black line). **D-F** The RMS amplitudes of the waveforms averaged across a two-minute-long trial are shown in the corresponding heatmaps. Some channels were removed due to high noise levels.

The heatmaps in Figure 3D-F represent the root-mean-square (RMS) amplitudes of the MUAP waveforms detected by spike-triggered averaging applied to each electrode in the sleeve array using the discharge times for the three motor units shown. Each heatmap contains a distinct hotspot representing the location in the array that detected the largest signal from each motor unit. These hotpots represent the innervation territory for each motor unit, which appears very focal for Index MU3 (Fig. 3E), broad for Index MU4 (Fig. 3F), and patchy for Index MU1 (Fig. 3D).

### Motor Neuron Activity in Intact and Paralyzed Muscles

The MUAPs provide a direct indication of the output of alpha motor neurons in the spinal cord. Rasters of motor neuron discharge events for the wrist and index tasks are shown in Fig. 4A-B. Many neurons, especially those associated with wrist movements (Fig. 4A), exhibited discrete bursts of activity corresponding to the flexion or extension phases of the task (Fig. 4A). To identify patterns of correlated firing among subpopulations of motor neurons, we performed non-negative matrix factorization on the smoothed firing rates (1-s smoothing window) for each motor neuron (Fig. 4C-D). For each task, two components, that we will refer to as neural modules, explained more than 80% of the variance in the original set of firing rates.

**Figure 4.**
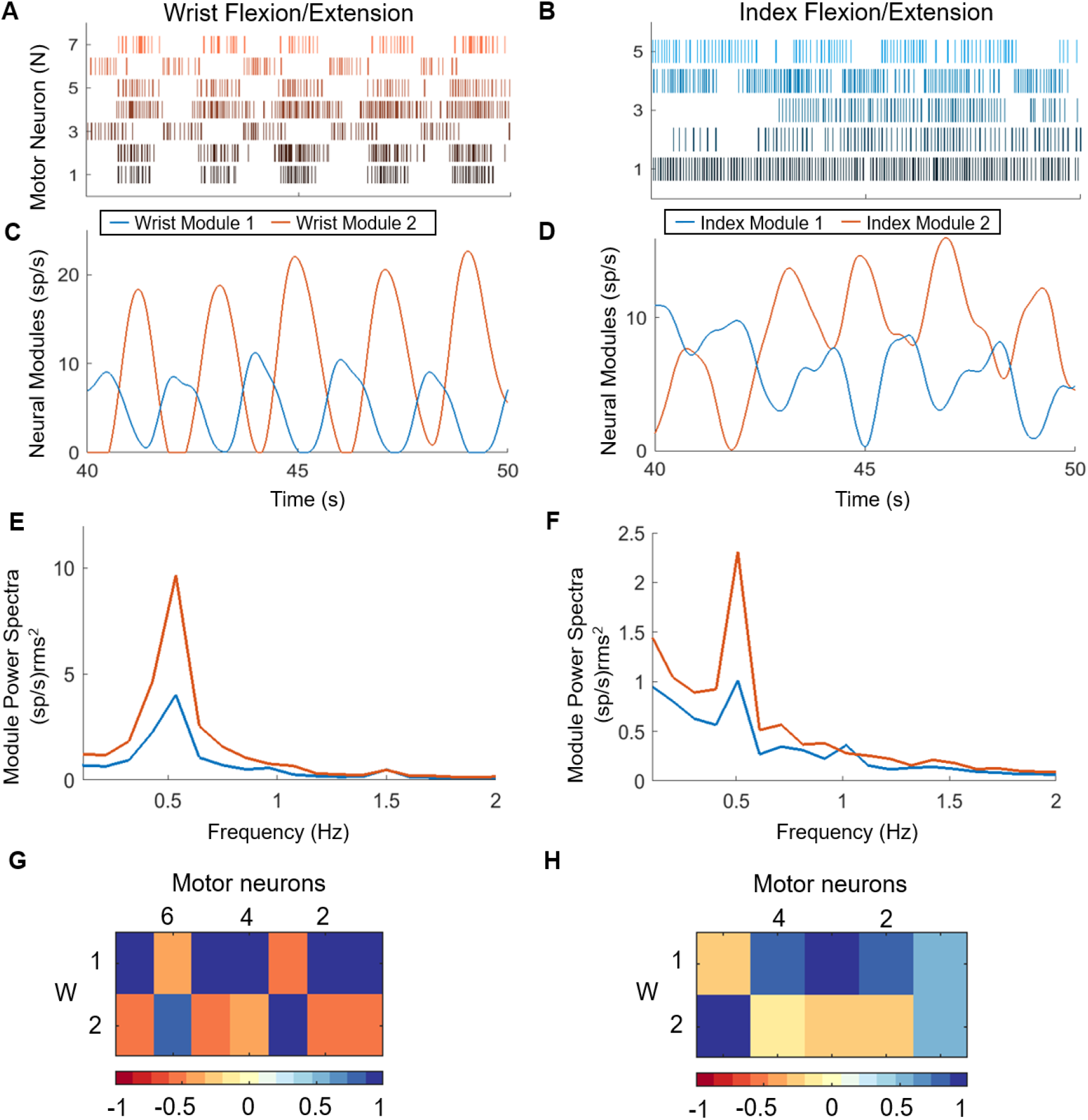
Extraction of neural modules from the discharge patterns in a population of motor neurons. Raster plot of motor neuron spiking during one block of 5 alternating flexion and extension movements of the wrist (**A**) and attempted movements of the index finger (**B**). Non-negative matrix factorization (NNMF) was applied to the smoothed firing rates of the motor neurons to generate 2 neural modules representing the composite firing rates of motor neurons that were active during the wrist (**C**) and index (**D**) tasks. Power spectra of the modules for the wrist (**E**) and index (**F**) demonstrates the phasic nature of the activity across all movement blocks at the 0.5 Hz cycle frequency. The bivariate correlation matrices between each of the motor neuron smoothed discharge patterns with the two components extracted by NNMF are shown in weight matrices (**G**, wrist; **H**, index).

Seven motor units were detected during the wrist task, and the 2 neural modules exhibited alternating phases of activity and quiescence (Fig. 4C), corresponding to the flexion and extension movements. Wrist Module 2 was associated with wrist extension and reached a peak firing rate approximately twice that of Wrist Module 1, which corresponds to wrist flexion. The asymmetry in peak firing rates agrees with the asymmetric pattern of weakness in the forearm muscles of this participant. MMT scores indicated that the participant could extend his wrist fully with gravity removed but not against gravity with resistance, and he had no visible movement of his wrist flexors.

Five motor units were detected during the index digit task, but the patterns of spiking activity for most neurons did not exhibit clear or alternating bursts of activity (Fig. 4B). The two neural modules for the index task show some rhythmic and alternating phases of activity. A 10 second interval of activity is shown in Figure 4D and reveals an interesting pattern in Index Module 2, with the first cycle exhibiting a normal pattern of activity and quiescence that was followed by subsequent cycles of activity appearing to ride on top of a tonic level of activity that persists and eventually fades. The patterns of spiking activity for Index Module 1 are less rhythmic, although some periodicity is evident.

The weight matrices in Figure 4 G-H show the relative contributions of each motor neuron to the 2 modules. During the wrist task, Wrist Module 1 corresponds to the extension phase and is dominated by 5 of the 7 motor units, while Wrist Module 2 comprises 2 units (motor neurons 3 and 5) that are active during the flexion phase. The composition of the neural modules for the index task is less distinct. Index Module 1 is dominated by 3 units (motor neurons 2, 3, and 4), and Index Module 2 comprises one dominant unit (motor neuron 5). Interestingly, motor neuron 1, contributes equally to both neural modules, which is understandable given the tonic pattern of activity that is evident in the spike rasters for that unit (Fig. 4B).

A Fourier transform was used to quantify the strength and frequency of task-related oscillations in each neural module. The power spectra across all movement blocks were averaged for the wrist and index trials (Fig. 4E-F). All neural modules exhibited a distinct peak in their power spectrum at the cycle frequency of the individual flexion and extension movements (0.5 Hz). The peak power for both wrist modules was more than 3 times larger than the peak power in the index modules, and in both tasks, the peak power for Wrist Module 2 (extension) was approximately twice as large as Wrist Module 1 (flexion). The relative weakness of the wrist flexor muscles, which scored 0 on manual muscle testing, is evident in the lower peak power observed in Wrist Module 1, which is associated with wrist flexion. The power spectra for the index neural modules exhibit the same asymmetry in the 2 motor modules as seen at the wrist.

Interspike interval (ISI) durations were extracted from the discharge times of each motor neuron. The ISIs across all motor neurons were below 200 ms and within observed physiological ranges (Fig. 5A-B). The ISIs of digit units (Fig. 5A) were comparable to those exhibited by units active during voluntary wrist and elbow movements (Fig. 5B). However, a couple of motor neurons had ISI distributions with large peaks, indicating a relatively higher level of activity (e.g. red distributions in the plots for the middle and index fingers).

**Figure 5.**
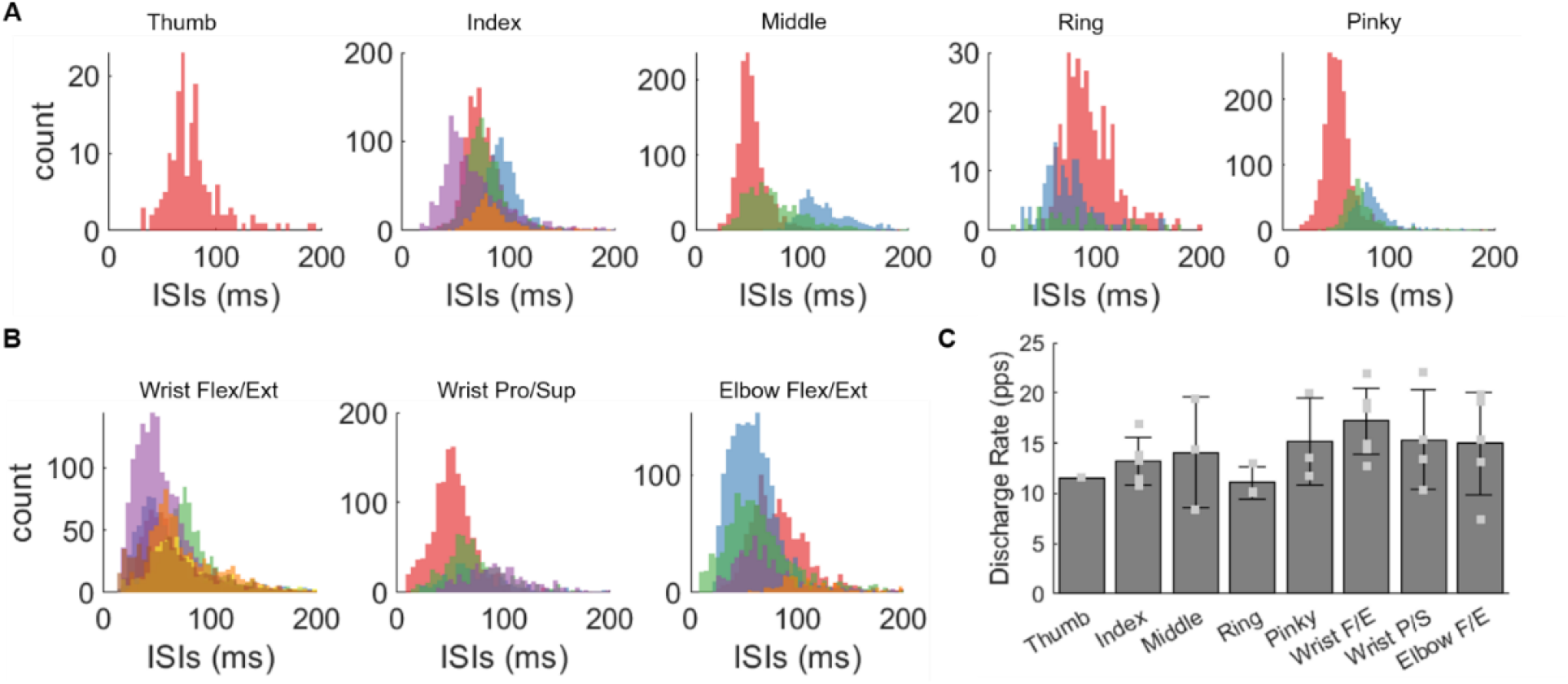
Motor unit properties across all tasks. Histograms of the interspike intervals (ISIs) for motor neurons (colors indicate different motor neurons) that were active during attempted movements of the digits (**A**) and during overt movements of the wrist (flexion/extension), forearm (pronation/supination), and elbow (flexion/extension) (**B**). **C** Bar plot showing the average motor unit discharge rate across the motor units associated with the movement task; error bars indicate the standard deviation

The discharge rates of all identified motor neurons were also within normal physiological ranges (Fig. 5C). We divided the motor neurons into two groups, those that were active during overt movement tasks (motive, n=16) and those associated with attempted movement (non-motive, n=15). The average discharge rate across motive neurons was 16.0 ± 4.0 (mean ± standard deviation) spikes per second (sp/s) and was significantly higher (p<0.05) than the average discharge rate of the non-motive neurons, which was 13.2 ± 3.2 sp/s. The standard deviation of discharge rates across the motive (8.9 ± 3.4 sp/s) and non-motive (4.9 ± 2.1 sp/s) neuron groups was also significantly different (p<0.05). Three motor neurons in the motive group were also active in the non-motive group as they were active during a subset of both digit (index, middle, and ring) and wrist/elbow tasks. These shared motor neurons fired at significantly higher rates during overt wrist and elbow movements (21.31 ± 1.26 sp/s) than during attempted digit movements (12.44 ± 3.90 sp/s) (p<0.05).

### Discriminating Task-Specific Activity in EMG and Motor Neuron Firing Rates

Task-specific variations in the spatial distribution of myoelectric activity were measured in the sleeve array (Fig. 2). To further examine task-specific differences in the spatial patterning of muscle activity, we quantified the degree to which the task-specific patterns of EMG and motor neuron firing rates could be discriminated using linear discriminant analysis (LDA) to classify activity across the five digits and the rest condition. Before classification, principal component analysis (PCA) was applied to the RMS EMG signals to reduce the dimensionality of the dataset to the first ten principal components, matching the number of motor units that were used for classification.

The median overall accuracy was 76% using the RMS EMG as input and 76.5% using the smoothed firing rates of the motor neurons as input (Fig. 6A-B). When using the RMS of the EMG, the mean individual task-classification accuracies ranged from 55% for the middle finger to 92% for the rest condition. The pinky (84%) and index fingers (79%) were classified with highest accuracy among the fingers (Fig. 6A). The errors were distributed across the digits and rest condition. When using motor unit firing rates for classification, the lowest individual accuracy was found for the thumb (35%), while the rest category was classified perfectly (100%) (Fig. 6B). The index finger was classified with the highest accuracy (89%) among the digits, followed by the pinky finger (83%) (Fig. 6B). The index task yielded the greatest number of motor units (n=5) of all digits (Table 1) and showed the strongest activity (Fig. 2). The thumb (35%) and ring (45%) fingers were classified poorly and most often confused with the rest category. Also, only a single unit was identified during the thumb task, and its activity was not strongly modulated with the cues. Overall, most errors in digit classification could be attributed to confusion with the rest condition (Fig 6B). These errors were due to the absence of motor unit activity during the movement blocks, resembling that of the rest condition.

**Figure 6.**
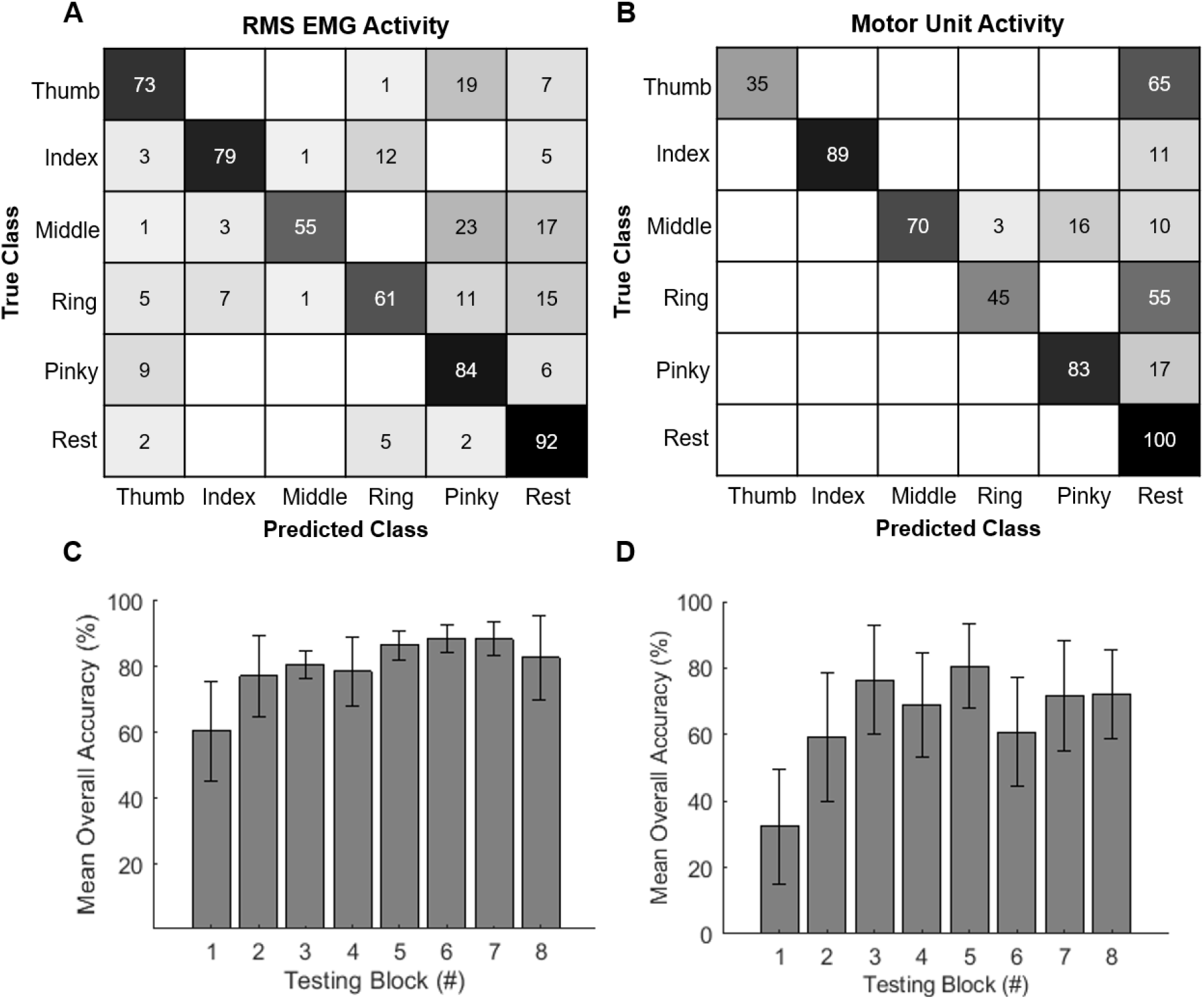
Offline classification using surface EMG or individual motor units. **A** The confusion matrix shows the overall individual classification accuracy across movements when the first ten principal components from the RMS EMG activity were used as input to an LDA classifier. The overall individual accuracy is the average accuracy for each finger across 8 testing sets. **B** The confusion matrix shows the overall classification accuracy across movements when the firing rates of ten identified motor units were used for classifier input into an LDA. **C-D** The mean overall accuracy is shown when each of eight movement blocks was used as the testing block and the remaining seven blocks were used for training. The error bars indicate the standard error across the individual digit accuracies.

Mean overall accuracy increased from the first to last movement block in both the RMS of the EMG and motor unit firing rate cases. In the EMG classification, the mean overall accuracy increased approximately 17% from the first to second block and approximately 28% from the first to sixth block (Fig. 6C). In the motor unit classification, the accuracy increased from 32.4% to 59.4% from the first to second block and reached a maximum of 80.6% in the fifth block (Fig. 6D).

We further tested the feasibility of implementing motor neuron decomposition and classification algorithms under conditions that simulated real-time operation. A playback of the recorded data was used to simulate a live data stream that was decomposed online with an average delay of 1.20 ms and with an accuracy comparable to the offline decomposition (96.0 ± 1.9% rate of agreement between online and offline decomposition) (Fig. 7). A binary classifier that was trained to recognize unique combinations of motor unit discharge patterns was used for online classification. Because this method required motor units to be firing in order to correctly classify the digit, only the time windows where a motor unit was active were included in the online classification. The unique combinations are shown in Table 2, where a 1 or 0 indicates whether the motor neuron (labelled a-j) was firing or not firing, respectively, during the specified condition. The classification accuracy was highest (100%) for the thumb, index, and pinky digits during periods when those units were active. For the middle and ring fingers, the classification accuracy was maximal only when the three identified neurons were firing concurrently, since the middle and ring fingers shared two neurons (see Table 2).

**Table 2.** Truth table used for detecting task-specific patterns of motor unit activity during simulated online decoder testing. Each row in the truth table indicates the activity state of each motor neuron (0 = not firing, 1 = firing) during attempted movements of each digit. Units a, b, c, e, h, and j were uniquely associated with one digit, while the others (d, f, g, and i) were associated with multiple digits. A unique combination of active motor units was used to detect movement of each digit.

|  | Activity State of Motor Units |  |  |  |  |  |  |  |  |  |
| --- | --- | --- | --- | --- | --- | --- | --- | --- | --- | --- |
| Units | a | b | c | d | e | f | g | h | i | j |
| Thumb | 0 | 0 | 0 | 0 | 0 | 0 | 0 | 1 | 0 | 0 |
| Index | 1 | 1 | 1 | 1 | 1 | 0 | 0 | 0 | 0 | 0 |
| Middle | 0 | 0 | 0 | 1 | 0 | 1 | 1 | 0 | 0 | 0 |
| Ring | 0 | 0 | 0 | 1 | 0 | 0 | 1 | 0 | 1 | 0 |
| Pinky | 0 | 0 | 0 | 0 | 0 | 1 | 0 | 0 | 1 | 1 |
| Rest | 0 | 0 | 0 | 0 | 0 | 0 | 0 | 0 | 0 | 0 |

**Figure 7.**
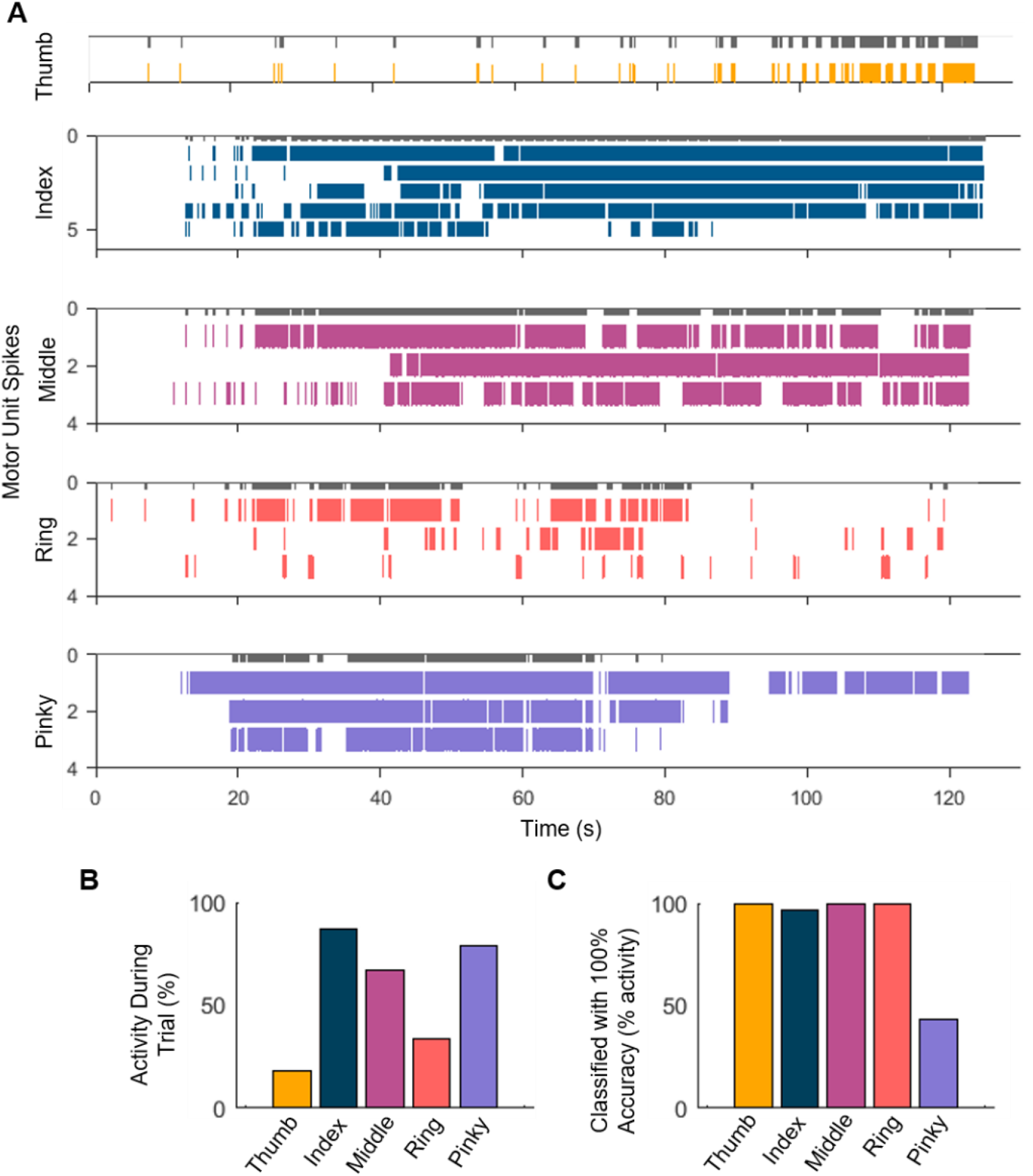
Simulated online implementation of motor unit decomposition and classification. **A** Raster plot of motor neuron spiking during the trials corresponding to attempted movements of each digit. We used the unique motor units or combination of multiple motor units to build a custom classifier to decode the activity of the individual fingers. The classifier was used to decode the attempted hand movements in 250 ms time frames. The grey bars on top of each spike raster depict successful classification.**B** The total activity of the motor neurons with respect to the trial length.**C** The percentage level of the classifier with respect to the maximum motor neuron activity across the individual hand digit tasks.

### High-Density Electromyography (HDEMG)

Previous studies of motor unit decomposition have used high-density EMG (HDEMG) electrode arrays with electrode diameters and center-to-center spacing <= 10 mm (19), both of which are much smaller than the size and spacing of electrodes in the sleeve array. Therefore, we performed an additional experiment to compare the properties of myoelectric signals measured with the sleeve array against those measured with commercially available HDEMG electrode arrays (TMSi, Inc.). In this experiment, a pair of HDEMG patch arrays was used, each comprising a 64-channel array of 4.5 mm diameter Ag/AgCl electrodes spaced 8.75 mm apart (center-to-center) arranged an 8×8 grid. One patch array was placed on each the dorsal and ventral surfaces of the forearm, approximately midway between the elbow and wrist (Fig. 8D). HDEMG was recorded while the participant attempted hand and wrist movements like those described above (Fig. 8D). Elbow movements were not included because the size and location of the HDEMG patch prohibited coverage of relevant muscles. The data presented here was obtained from the patch placed over the extensor muscles (Fig. 8D, lower panel). The dataset using the HDEMG patch was collected approximately 18 months after the sleeve array experiments. The HDEMG data provides a benchmark for comparing the quality (i.e. signal amplitude and noise) of EMG and motor unit data recorded by the sleeve.

**Figure 8.**
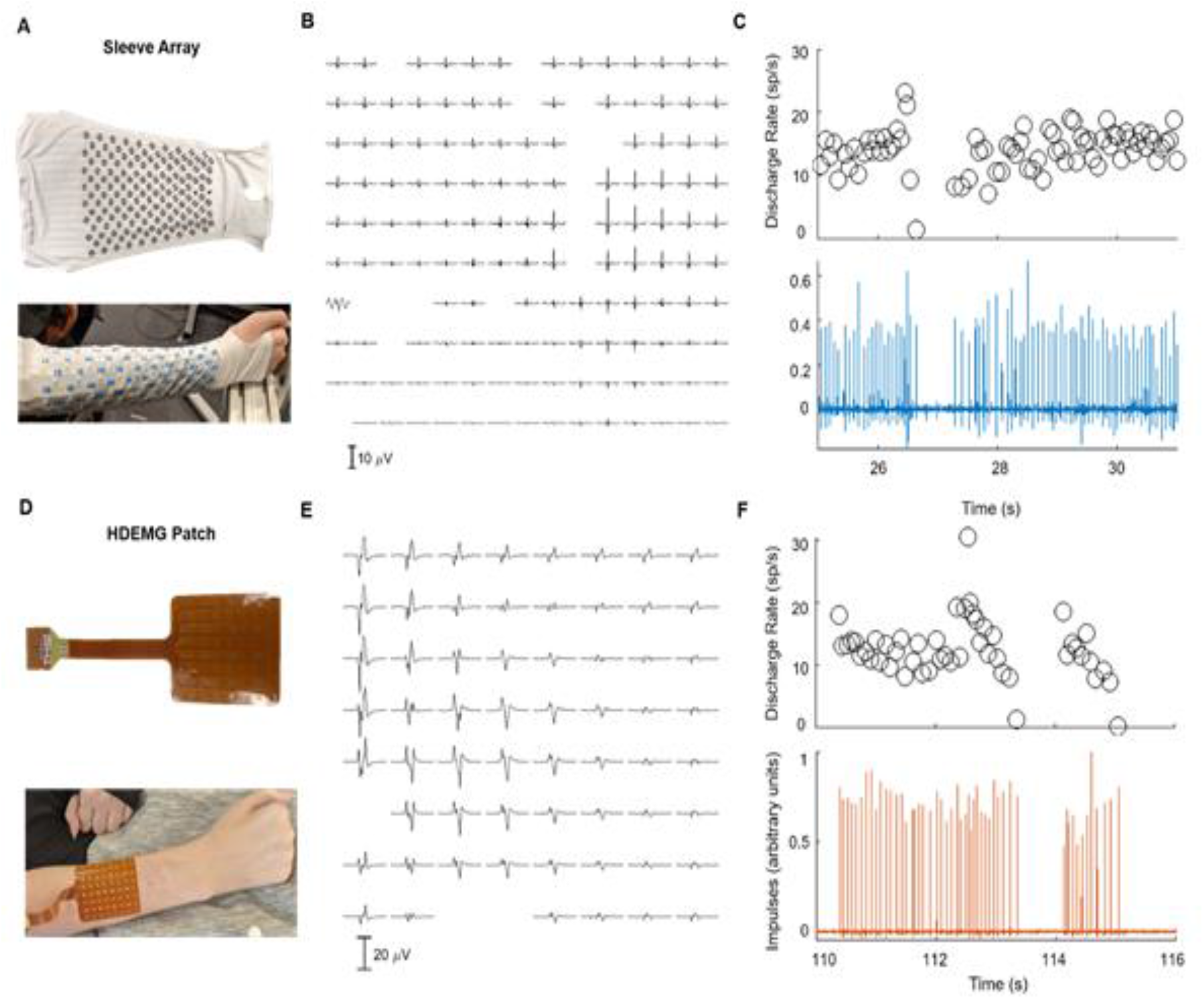
Comparison of recordings by commercial HDEMG patch and the wearable sleeve array. **A** Wearable sleeve array consisting of 150 electrodes (12 mm diameter, 25 mm inter-electrode distance) that covers most of the activity from the forearm muscles. **B** The spike-triggered average motor unit action potential over the 150 electrodes. Note that some channels are not shown because of noise. **C** The discharge activity for one representative motor unit with the corresponding activity index (lower panel) extracted by blind source separation.**D** Commercially-available HDEMG patch consisting of 64 electrodes (4.5 mm diameter, 8.75 mm inter-electrode distance) over the forearm extensor muscles. **E-F** The average spike-triggered motor unit action potential and the discharge activity of one representative motor unit (as shown in **B** and **C**).

Figure 8E shows an example of average MUAP waveforms detected on the dorsal HDEMG patch. The peak to peak amplitude of the MUAP waveform on many electrodes exceeds 20 μV. For comparison, Figure 8B shows a similar map of MUAP average waveforms for a unit detected on the sleeve array, which also reveals that the MUAP waveforms are detectable across many electrodes in the array. The motor neuron discharge rates for the motor units detected in the sleeve and HDEMG patch arrays are shown in Figure 8C,F, respectively, and are consistent with the normal physiological range of 10-20 sp/sec.

A total of 26 active motor units were identified across all tasks using the HDEMG patch (Table 1). Between two (index flexion/extension) and six (wrist flexion/extension) units were active during each task, and these units showed similar PNRs across tasks (Table 1). Notably, the average PNR across units using the HDEMG patch was generally higher than the average PNR for units detected on the sleeve array. The noise floor or baseline voltage levels recorded with the patch array were approximately five times lower (10.5 ± 4.6 μV_RMS_) than for the sleeve array (47.6 ± 16.8 μV_RMS_) (Table 1). Additionally, the average amplitude of the MUAP waveforms detected on the HDEMG patch were approximately three times lower (169.3 ± 97.4 μV) in comparison to the sleeve array (474.0 ± 51.9 μV) (Table 1).

## Discussion

Recovery of hand function after SCI is a top priority for people with tetraplegia (3, 4) and has motivated the development of implantable and wearable devices that reanimate or assist hand functions (5–12, 20). Brain computer interface (BCI) and EMG-based control interfaces are being developed to provide users with direct voluntary control of FES, robotic exoskeletons, and other devices for restoring hand function. While EMG-based control is used routinely by people with amputations to operate motorized prosthetic hands (21), the development of BCIs for assistive technologies has been motivated in part by the assumption that reliable control signals cannot be obtained readily from a paralyzed limb.

Recent studies have demonstrated that EMG signals can be measured in muscles that are weakened or even paralyzed by SCI (9–11). Thus, measuring and decoding myoelectric signals from severely impaired muscles may provide a practical, noninvasive solution for controlling assistive devices after SCI. Current systems, however, tend to utilize traditional bipolar EMG electrodes, which are bulky and require targeted placement over specific muscles. Furthermore, the power of EMG signals recorded from muscles below the level of SCI tends to be very small, resulting in poor signal-to-noise ratio and low accuracy control. We addressed both problems by creating a system comprising a novel wearable sensor array and signal processing methods that permit detection of motor unit action potentials. The firing rates of ∼20 motor neurons were measured simultaneously, exhibiting spatiotemporal patterning that was highly task specific.

We have shown that low-amplitude yet distinguishable muscle activity can be detected from paralyzed muscles during attempted digit movements, even in the absence of joint movement (Fig. 2). As expected, attempted movements of all fingers yielded considerably lower amplitude EMG (RMS) signals than overt movements of the wrist (Fig. 2). The RMS of the EMG signals was used to classify movements across the five digits offline, resulting in a median overall accuracy of 76%, while the firing rates of the identified motor neurons classified movements with a median overall accuracy of 76.5%. We have also shown in a simulated online demonstration that motor unit spiking activity may provide an ideal control signal in neuroprosthetic applications, correctly predicting movement intention 88 ± 24% of the time. LDA was used for offline classification to compare the information content of the RMS EMG and motor unit firing rates, while a binary classifier was used to simulate online classification to detect attempted movement of specific digits. The binary classifier is specifically suited to the type of information carried by motor unit spike trains.

Errors in classification could be attributed primarily to confusion between weaker EMG in certain task conditions (ring and pinky fingers) and the rest condition. Furthermore, we attempted to discriminate between attempted movements of individual fingers, which is a challenging task even for able-bodied subjects (22). In this study, the participant was only provided cues to instruct the type and timing of movements. EMG biofeedback was not provided, and the subject was unable to generate any movement in his fingers, making it impossible for him to evaluate his performance during the task. In the absence of feedback, the subject was unable to achieve consistent levels of recruitment and long periods of inactivity were observed in some cases (e.g. thumb, ring, pinky). However, during those periods when the subject generated motor unit activity during attempted movements, the patterns of motor unit activity were very distinct for each task.

Since the SNR is generally low for myoelectric signals generated by paralyzed muscles, we implemented a feature extraction method that maximizes the amount of information extracted from the sensor array. This method, motor unit decomposition, allowed us to extract the discharge timings of 20 unique motor units across attempted movements of the digits and overt movements of the wrist and elbow. Motor unit activity is a direct indicator of the spiking output of motor neurons in the spinal cord. Because motor units exhibit stereotypical recruitment properties (23), they may be the ideal control signal for discriminating between volitional and pathological signals, allowing for decoding of movement intention to be made more robust to abnormalities in the activity related to SCI, such as spasticity (24, 25).

To our knowledge, this represents the first demonstration of movement-decoding from motor unit firing rates after SCI. This method has the potential to enable researchers and clinicians to interface non-invasively with motor neurons exiting the lesioned spinal cord, creating new opportunities for controlling assistive devices, evaluating spared function after injury, and tracking recovery of neuromotor function during rehabilitation.

### Measuring and Tracking Changes in Motor Function

Wearable sensors offer the potential to not only aid in restoring hand function but also to provide an indicator of spared and recovered function after injury. We have demonstrated that measures of motor unit function, such as discharge rate, ISI, and amplitude, can be obtained from low levels of EMG recorded non-invasively. Distinct patterns or abnormalities in motor unit behavior can be used to detect pathologies, like spasticity (24, 25) and tremor (26, 27). In our results, the tonic activity present in Index Module 2 (Fig. 4D), representing attempted extension, may be related to abnormal physiology of the motor neuron after injury (28).

In addition to measures at the single unit level, global patterns of EMG activity and the number of active motor units may also be beneficial in monitoring recovery after injury. If tracked over time, these measures could provide a sensitive metric of adaptation and recovery in the spinal cord following injury. This wearable system would allow clinicians to monitor treatment and rehabilitation protocols over time and tailor them as necessary. This technology could also be useful in evaluating arm and hand function in a range of populations including those with Parkinson’s disease, amyotrophic lateral sclerosis (ALS), and stroke. The simple setup and comfort of the sleeve array would allow it to be used during inpatient or outpatient therapy as well as during daily activities within the home.

## Conclusions

SCI at the cervical level disrupts connections between the brain and torso and limbs, often leaving individuals with little or no control over movement of both hands. Hand function is not only vital to performing activities within the home but also likely plays a key role in whether individuals return to employment after injury. It has been shown that individuals with more severe injuries are less likely to become employed after injury (29). Therefore, individuals with tetraplegia have ranked arm and hand function as the most important area to recover among lost functions. Researchers have attempted to restore hand and arm control by providing individuals with control over various assistive technologies. Our method utilizes a wearable sleeve array that detects low levels of myoelectric activity and allows for the extraction of spinal motor neuron activity. The motor neuron activity can be leveraged to control assistive devices or as an indicator of function after injury. This system has the potential to allow individuals with SCI to live more independently by restoring intuitive hand and arm control.

## Materials and Methods

### Participant

One participant with tetraplegia was involved in this study. Data was collected as part of an ongoing intracortical brain-computer interface trial being conducted under an FDA Investigational Device Exemption and approved by the University of Pittsburgh Institutional Review Board (NCT01894802). Informed consent was obtained from the participant prior to any experimental procedures. Intracortical data were not used in the analyses presented here.

The participant was a male between 30 and 35 years old with a C5 motor and C6 sensory American Spinal Injury ASIA Impairment Scale B (motor complete/sensory incomplete) spinal cord injury sustained 14 years prior to the experiment. A Manual Muscle Test (MMT) was performed to assess the strength and function of the participant’s wrist flexors and extensors and finger flexors and extensors (16). The participant was able to extend his wrist fully with gravity removed but not against gravity with resistance (MMT score, 4-). He had no visible movement of his wrist flexors, finger flexors, or finger extensors (MMT scores, 0); however, low levels of myoelectric activity were detected from his right forearm.

### Experimental Setup

The participant was seated in his wheelchair facing a computer monitor and with his right arm placed on his lap. A sleeve array (Battelle Memorial Institute, Columbus, OH; Fig 8A) was placed on the right arm of the participant. The sleeve array comprises a stretchable fabric with an embedded array of 150 stainless steel disc electrodes (12 mm diameter) spaced approximately 25 mm apart (center-to-center) and spanning the full circumference of the forearm from elbow to wrist. The sleeve has a zipper along the ulnar edge of the forearm to provide closure. Prior to donning the sleeve, the participant’s forearm was cleaned and a conductive cream (Saebo, Charlotte, NC) was applied to the skin. Impedance measurements were measured at 1 kHz before beginning recording. Monopolar EMG signals were sampled at 30 kHz using an Intan Recording Controller (Intan Technologies, Los Angeles, CA). Fourteen channels of the 150-channel sleeve array were removed before further processing due to high levels of noise.

The participant was instructed to attempt a series of movements involving the finger digits, wrist, elbow, and shoulder. The experimenter explained to the participant to attempt the movements as if he were able to move his hand as opposed to imagining performing the instructed movements. A video was shown to cue the movements, where an able-bodied person demonstrated each task. The participant was instructed to follow the video and attempt to match each movement shown. Movement trial types included flexion/extension of each finger digit, flexion/extension of the wrist, pronation/supination of the wrist, flexion/extension of the elbow, and flexion/extension of the shoulder. Each trial consisted of 8 blocks of 5 alternating flexion and extension movements (Fig 1E). Opposing movement cues (e.g. flexion/extension or pronation/supination) were alternated at a rate of one per second.

Data were also collected during a second session with the same participant 18 months after the original session. In this second session, two 64-channel HDEMG path arrays were used to detect EMG data from a subset of the extensor and flexor muscles in the forearm (TMSi, Netherlands; Fig 8D). One array each was applied over the extensors and flexors. Each array comprises an 8×8 grid of Ag/AgCl electrodes having a diameter of 4.5 mm and are spaced 8.75 mm apart. As in the sleeve array setup, an implanted skull screw was used as the ground electrode for EMG. Monopolar EMG signals were sampled at 4 kHz using a TMSi SAGA 64+ recording system (TMSi, Netherlands). The participant was given video instruction to attempt the same set of movements, excluding movements of the elbow and shoulder and including cylindrical grasp and hand extension movements.

### Motor Unit Decomposition and Analysis

The surface EMG signals were decomposed into constituent motor unit action potentials (MUAPs) by blind source separation in offline (18, 30) and online (31) implementations. We used a convolution kernel compensation technique that has been described previously (17) and validated in synthetic datasets (18, 30), intramuscular EMG motor unit recordings (32, 33), and during a wide-range of voluntary force contractions in both healthy humans (34, 35) Parkinson’s disease (36) and after targeted muscle reinnervation (37, 38). Briefly, the surface EMG is a convolutive mixture of MUAPs. The decomposition technique utilizes the high spatial dimensionality of the MUAP provided by the high-density EMG recordings to isolate the unique MUAP waveforms.

From the motor unit discharge timings, we extracted the two-dimensional action potential waveforms by spike-triggered averaging of the firing instances of each motor neuron. The high-density EMG signals were decomposed for each contraction individually. After blind source separation, we used the two-dimensional action potential as a spatial filter to manually check the reliability of each discharge for all motor units. Only motor units that showed a >26 pulse-to-noise ratio (PNR, dB) were manually inspected (18). The PNR represents an estimate of the signal to noise ratio of the extracted motor units and thus the accuracy of decomposition. We have reported previously that motor units with PNR >26 show high validity (18) and reliability (35) across a wide range of voluntary force intensities.

We then tested the accuracy in decomposition by computing the two-dimensional cross-correlation function between a random selection of MUAPs, as recently done in neurologically healthy individuals (35). This procedure correlates the motor unit action potential between different time frames of the discharge timings of the motor neuron.

The motor unit decomposition procedure was applied separately for each task. To detect motor units that were active during multiple tasks, we verified the uniqueness of each MUAP waveform by two-dimensional cross-correlation across all identified motor neurons. Therefore, the two-dimensional cross-correlation represents an estimate of the similarity in waveform shape across all action potentials within the sleeve array.

Further, we performed two-dimensional correlations between all possible motor unit pairs within and between the different contractions to track repeatedly-active motor units across task conditions. We will refer to this procedure as the motor unit tracking technique. Briefly, the motor unit waveforms were extracted in 15 ms time windows with spike-triggered averaging. The two-dimensional waveforms were cross-correlated across all channels of the sleeve array at different time frames. The two-dimensional cross-correlation represents the two-dimensional version of a one-dimensional cross-correlation, therefore ranging between −1 and 1, with 1 if the motor unit action potential is the same.

After testing the validity of the discharge timings for the motor units, we computed several neurophysiological estimates. The motor unit interspike intervals (ISIs) correspond to the time difference between two discharge timings of a motor unit. Absolute values of the instantaneous firing rates were computed as the inverse of the ISI. Moreover, we performed a non-negative matrix factorization (NNMF) of the smoothed motor unit discharge timings. For this purpose, a convolution of the motor neuron spike instants (binary signal) with a 1-s Hanning window extracted the smoothed discharge rates for each motor neuron. The non-negative matrix factorization (100 iterations), extracted the common components within a population of motor neurons during each attempted task. The number of factors was established by computing the variance accounted for (reconstruction accuracy) with respect to the extracted neural modules in the pool, in a similar way as done in muscle synergy studies (39). For all cases, we identified two factors explaining most of the variance of the discharge patterns of the motor neurons (see results). The discharge timings for NNMF were smoothed with a 1-s time window since this window size approximates the muscular dynamics, which have a low frequency bandwidth (<2Hz).

Finally, coherence analysis was performed on all combinations of groups of randomly permuted motor units for each trial. Coherence was estimated with Welch’s averaged periodogram with non-overlapping Hanning window of 1-s duration.

### Classification

All EMG signal processing was performed using MATLAB (Mathworks Inc., Natick, MA). The monopolar signals were band-pass filtered in the 20-450 Hz range (4th order, zero-lag Butterworth) (Fig. 1D). Before further analysis, selected channels (14 total) were removed from all trials due to high levels of baseline noise, leaving 136 channels of monopolar EMG for the analysis. A sliding window method was used to calculate the root-mean-square (RMS) amplitude of the bandpass-filtered EMG in 200 ms bins with 50% overlap.

Motor unit discharge rates were calculated as the inverse of the ISIs and smoothed using a Gaussian distribution in 50 ms bins across each trial. If a motor unit was not active during a specific trial, the corresponding smoothed firing rate values were set to zero. Only activity recorded during finger movement trials was used in this analysis in an effort to determine the discriminability of attempted movements from clinically paralyzed muscles.

Principal component analysis (PCA) was performed to reduce the dimensionality of the surface EMG dataset. Data collected during finger movement trials (thumb, index, middle, ring, and pinky digits) were aggregated before applying PCA. PCA was performed across the RMS of the EMG activity of all electrode channels (n=136). After applying PCA, the time series corresponding to the first ten principal components representing the surface EMG activity were selected for classification by linear discriminant analysis (LDA). All ten identified motor units were also used for a separate classification by motor unit firing rates. LDA was chosen to classify digit movement as it has been shown to be a simple, yet effective option for pattern classification of myoelectric activity (40). For both surface EMG and motor unit classification, a single block, composed of 10 movements, from each trial was used for cross-validation, totaling 50 individual movements per test set. The remaining seven blocks in each trial were compiled and used for training. Accuracy was calculated as the total number of correct movement labels divided by the total number of labels in the testing set. The mean individual digit accuracies, averaged over all testing sets, are reported in the diagonal blocks of the confusion matrices (Fig. 6A-B). Overall classification accuracy was calculated by averaging the mean individual accuracies.

The simulated online classification software was based on the labelling of the motor units, as shown in Table 2. First, agonist/antagonist weights were extracted from 10 seconds of training data using non-negative matrix factorization. The greatest weights were then binarized for each motor unit corresponding to either agonist and antagonist activity. The motor units that were identified as “unique” (not present in any other trials) or the combination of unique motor units were used as triggers to classify the hand digits. Motor unit spike instants were then filtered with a 1-Hz window convolution, which represents the bandwidth of the neural drive to the muscle that is translated into force. Motor unit labelling was performed offline and then implemented in the online decoder as a binary classifier. Table 2 shows the activity state of the motor units (labelled a-j), where a 1 specifies an active, or firing unit, and a 0 represents an inactive unit. Specifically, each finger was associated with a unique motor unit discharge pattern (representing either a unique motor unit or unique combination of motor units for a specific finger) in a 250 ms window. If the unique motor unit discharge patterns in a 250 ms time frame corresponded to the tested finger, it was correctly classified. Because some motor units were shared between conditions, we computed the classification accuracy with respect to the total active time across conditions (Fig. 7A-C, Table 2). The effectiveness of this method required motor units to be firing to correctly classify a specified digit. Therefore, only the 250 ms time frames where a motor unit was spiking were included in the online classification.

### Statistical Analysis

Blind source separation was utilized to decompose surface EMG signals into constituent motor unit action potentials (MUAPs). Motor unit decomposition was applied to the surface EMG recordings using a validated convolution kernel compensation technique to obtain the discharge timings of active motor units (17). Two-dimensional action potential waveforms were extracted from the motor unit discharge timings by spike-triggered averaging. These two-dimensional action potentials were used as a spatial filter to check the consistency of the spike firing for all motor units. Extracted motor units with a PNR >26 were manually inspected to determine their validity (18). Decomposition accuracy was verified by computing the two-dimensional cross-correlation function between random selection of MUAPs (35). A motor unit tracking procedure was performed to track motor units that were active consistently across different tasks. Two-dimensional MUAP waveforms were cross-correlated across all channels of the array at varying time points; resulting values ranged from −1 to 1, where one indicates that the MUAPs are the same.

Motor unit properties are reported as the mean and standard deviation across all units active during a particular task (Table 1). The differences in mean and variance of the discharge rates associated with the motive and non-motive motor neuron groups were tested with a two-sample *t*-test with significance level of 0.05. Classification of the digits during attempted flexion and extension movements was performed using LDA and implemented in MATLAB. The classifier was trained using seven of eight blocks from each trial and tested using the remaining block from each trial. These training and testing procedures were repeated until each of the eight total blocks had been used for testing. Individual digit accuracies were averaged over all testing sets, and the mean values are shown in the diagonal blocks of the confusion matrices in panels C and D of Fig. 6. The median overall accuracy was calculated by taking the median of the mean individual accuracies.

## Data Availability

Anonymized data created for the study are or will be available upon request.

## Acknowledgements

The authors would like to thank the participant for his dedication to the study and Dylan Royston for contributing the instructional videos used during data collection. Research reported in this publication was supported by the National Institute Of Neurological Disorders And Stroke of the National Institutes of Health under Award Numbers UH3NS107714 and U01NS108922 and Defense Advanced Research Projects Agency (DARPA) and Space and Naval Warfare Systems Center Pacific (SSC Pacific) under Contract N66001-16-C4051. The content is solely the responsibility of the authors and does not necessarily represent the official views of the National Institutes of Health, DARPA, or SSC Pacific. JT and DW were supported by a National Science Foundation Award, #CHS1717654. AdV and DF are partly supported by the European Research Council Synergy project Natural BionicS, #810346.

